# A complex ePrescribing-based Anti-Microbial Stewardship (ePAMS+) intervention for hospitals combining technological and behavioural components: protocol for a feasibility trial

**DOI:** 10.1101/2022.04.27.22274394

**Authors:** Christopher J Weir, Imad Adamestam, Rona Sharp, Holly Ennis, Andrew Heed, Robin Williams, Kathrin Cresswell, Omara Dogar, Sarah Pontefract, Jamie Coleman, Richard Lilford, Neil Watson, Ann Slee, Antony Chuter, Jillian Beggs, Sarah Slight, James Mason, Lucy Yardley, Aziz Sheikh

**Affiliations:** Edinburgh Clinical Trials Unit, Usher Institute, University of Edinburgh, Edinburgh, UK; Usher Institute, University of Edinburgh, Edinburgh, UK; Newcastle upon Tyne Hospitals NHS Foundation Trust, Newcastle, UK; Institute for the Study of Science, Technology and Innovation, University of Edinburgh, Edinburgh, UK; Department of Health Sciences, University of York, York, UK; Institute of Clinical Sciences, University of Birmingham, Birmingham, UK; University of Birmingham, Birmingham, UK; NHS Covid Vaccine North East and North Cumbria, Chief Pharmacist, Newcastle upon Tyne Hospitals NHS Foundation Trust, Newcastle, UK; NHS England, UK; School of Pharmacy, Newcastle University, Newcastle, UK; Warwick Medical School, University of Warwick, UK; School of Psychological Science, University of Bristol, Bristol, UK; School of Psychology, University of Southampton, Southampton, UK

## Abstract

**Introduction:** Antimicrobial resistance is a leading global public health threat, with inappropriate use of antimicrobials in healthcare contributing to its development. Given this urgent need, we developed a complex ePrescribing-based Anti-Microbial Stewardship intervention (ePAMS+).

**Methods and analysis:** ePAMS+ includes educational and organisational behavioural elements, plus guideline-based clinical decision support to aid optimal antimicrobial use in hospital inpatients. ePAMS+ particularly focuses on prompt initiation of antimicrobials, followed by early review once test results are available to facilitate informed decision-making on stopping or switching where appropriate. A mixed-methods feasibility trial of ePAMS+ will take place in two NHS acute hospital care organisations. Qualitative staff interviews and observation of practice will respectively gather staff views on the technical component of ePAMS+ and information on their use of ePAMS+ in routine work. Focus groups will elicit staff and patient views on ePAMS+; one-to-one interviews will discuss antimicrobial stewardship with staff and will record patient experiences of receiving antibiotics and their thoughts on inappropriate prescribing. Qualitative data will be analysed thematically. Fidelity Index development will enable enactment of ePAMS+ to be measure objectively in a subsequent trial assessing the effectiveness of ePAMS+. Quantitative data collection will determine the feasibility of extracting data and deriving key summaries of antimicrobial prescribing; we will quantify variability in the primary outcome, number of antibiotic defined daily doses (DDD), to inform the future larger-scale trial design.

**Ethics and dissemination:** The qualitative research and Fidelity Index were approved by the Health and Research Authority and the North of Scotland Research Ethics Service (ref:19/NS/0174). The feasibility trial and quantitative analysis were approved by the London South East Research Ethics Committee (ref:22/LO/0204). Findings will be shared with study sites and with qualitative research participants and will be published in peer-reviewed journals and presented at academic conferences.

**Trial registration:** ISRCTN 13429325 (protocol v1.0, 15/12/2021)

**Strengths and limitations:**

- Mixed-methods study, incorporating qualitative and quantitative elements, assessing feasibility of a trial evaluating the ePrescribing-based Anti-Microbial Stewardship (ePAMS+) intervention.
- The feasibility trial will inform refinements of ePAMS+ intervention and its future full-scale evaluation.
- Development of a Fidelity Index to enable adherence to the ePAMS+ intervention to be assessed objectively.
- Two study sites may limit generalisability, although inclusion of several ward types will ensure the trial covers a breadth of clinical contexts.
- Implementation of ePAMS+ in the Cerner ePrescribing and Medicines Administration (EPMA) system means feasibility in other systems will still need to be established.

**Lay summary:** Not all infections are caused by bacteria. For those that are, antibiotics may be a suitable treatment. When patients first come to hospital, it is sometimes not clear what is causing their illness so doctors may prescribe antibiotics just in case until results from tests to identify the presence of microbes are available. The more antibiotics a person takes the more likely they are in the future to develop bacteria in their body that antibiotics are less effective at treating. The ePrescribing-based Anti-Microbial Stewardship (ePAMS+) intervention is designed to guide the appropriate use of antibiotics. ePAMS+ uses the hospital electronic patient health record to alert prescribers to situations where changing or stopping antibiotics may be a good option for a patient, consistent with existing national guidelines.

The ePAMS+ intervention will prompt healthcare professionals responsible for prescribing to review the progress and test results of a patient receiving antibiotics. After such a review:

- doctors may decide that a patient will need to carry on with antibiotics because they are right for their illness;
- healthcare staff may receive test results that can inform how long antibiotics should be prescribed for and which are best to treat the infection;
- patients may have their antibiotics stopped if the prompts alert the prescriber to decide that the illness is not caused by bacteria.

By implementing the ePAMS+ intervention in two hospitals and interviewing staff and patients, this study will assess whether ePAMS+ and our implementation methods are acceptable. It will also confirm whether it is possible to gather the data needed to assess how well ePAMS+ works. This will help design a future larger-scale study.

## Introduction

Antimicrobial resistance (AMR) has been highlighted by the World Health Organization (WHO) as one of the top 10 global public health threats facing humanity.^1^ In the European Union, antimicrobial resistant infections are estimated to be responsible for at least 25,000 deaths annually.^2^ Globally, these infections claim around 700,000 lives each year.^3^ Inappropriate and suboptimal use of antimicrobials in healthcare are key contributors to AMR,^4^ which can lead to an increase in and spread of resistant bacteria and increase risk of poor outcomes from bacterial infections due to a reduced number of effective antimicrobial therapeutics. It is therefore imperative to stem inappropriate antimicrobial use.^5^

The European Centre for Disease Prevention and Control revealed that the United Kingdom (UK) had the third highest hospital consumption of systemic antibiotics per capita in Europe,^6^ with hospital inpatient antibiotic consumption increasing by 6.3% between 2016-19.^7^ The English Surveillance Programme for Antimicrobial Utilisation and Resistance Oversight (ESPAUR) found increases in the rate of bloodstream infections caused by Escherichia coli and Klebsiella pneumoniae between 2016 and 2019,^7^ as well as a slight increase in the proportion of bloodstream infections resistant to piperacillin/tazobactam between 2016 and 2020. This increased resistance places further pressure on clinicians to use ‘last resort’ antibiotics such as carbapenems. The exceptional impact of the first waves of the severe acute respiratory syndrome coronavirus 2 (SARS-CoV-2) pandemic^7^ compounded these challenges, the 10.6% increase in hospital inpatient antibiotic consumption in 2019-20 potentially leading to increased inappropriate use.

In response to this growing threat, Public Health England (now UK Health Security Agency) championed guidance encouraging clinicians to “Start Smart–Then Focus” in relation to the initiation and maintenance of antibiotics.^8 9^ Moreover, the NHS England (NHSE) Antimicrobial Resistance and Antimicrobial Stewardship Commissioning for Quality and Innovation (CQUIN) aims to promote a “reduction in antibiotic consumption per 1,000 admissions”.^10^

As NHSE rapidly moves towards increasing digitisation of hospitals,^10^ electronic prescribing (ePrescribing) systems are crucial to antimicrobial stewardship (AMS) relating to prescribing.^11^ Furthermore, guideline-based clinical decision support (CDS) systems can help, and the effects of CDS rules can be enhanced through techniques that support clinicians and hospitals to prioritise AMS through, for example, facilitating timely review of antibiotics.^12^ A review exploring the appropriate use of antibiotics through hospital ePrescribing systems^12^ and related conceptual work^13^ indicate ePrescribing systems – integrated with behavioural and organisational support – have a major role in improving AMS. We have carefully conceptualised a complex ePrescribing-based Anti-Microbial Stewardship (ePAMS+) intervention that aligns with the UK government Five Year Antimicrobial Resistance Strategy^14^ and ESPAUR^7^.

The overall aim of our mixed-methods research programme is to plan, develop and optimise the ePAMS+ complex intervention and to assess its clinical and cost-effectiveness within a hybrid cluster-randomised stepped-wedge clinical trial. Prior to finalising the protocol of the full-scale trial assessing intervention effectiveness, we plan a feasibility trial involving testing of data extraction and the implementation and acceptability of the ePAMS+ intervention to inform the trial design.

### Aims

Our primary aims are to:

1. Explore user acceptability of the content of the ePAMS+ technical component and identify any barriers to use
2. Assess whether ePAMS+ Antibiotic Order Plans are used as intended in clinical practice and if not, identify barriers
3. Assess acceptability to healthcare professionals of the content of the ePAMS+ intervention plan (ePAMS+ organisational component) and training materials (ePAMS+ educational component)
4. Determine between-patient variability in total antibiotic consumption to enable planning of the full-scale cluster-randomised hybrid stepped wedge clinical trial
5. Develop processes of collecting outcome data from ePrescribing systems prior to and following introduction of ePAMS+.

Our secondary aims are to:

1. Understand how ePAMS+ may be best delivered across multiple care settings and site information systems
2. Determine whether the procedures for implementing ePAMS+ are acceptable and feasible
3. Assess whether ePAMS+ can be successfully integrated into hospital settings to enable changes in prescribing behaviour
4. Develop a Fidelity Index to quantify the extent to which core principles of ePAMS+ are enacted in antibiotic prescribing practice and test its usability
5. Confirm hypothesised mechanisms of action, refine programme theory and identify appropriate process analysis measures of mechanisms of action for a future full-scale trial.

## Methods and analysis

### Design

This initial phase aims to assess the feasibility of embedding the ePAMS+ intervention into existing technological systems and organisational practices and extracting trial outcome measures. The feasibility trial will be conducted between April and August 2022 within selected hospital departments at two NHS acute hospital provider organisations in England. It has three main elements:

1. Through focus groups, interviews and observation of practice, qualitative research will explore how the ePAMS+ intervention is received and how it may need to be adapted for other contexts. It will also identify likely mechanisms of action to be examined further in the follow-on process evaluation of the planned full-scale stepped-wedge trial evaluating the ePAMS+ intervention. We will seek to understand barriers and facilitators to implementation, including usability and acceptance issues.
2. Quantitative analysis will develop methods of deriving key summaries of antimicrobial use and will estimate the variability in these measures, using routine administrative data extracted from the Cerner ePrescribing and Medicines Administration (EPMA) system at each site.
3. We will develop a Fidelity Index prototype to assess the enactment of the ePAMS+ intervention. This will involve examining how closely practice matches its underlying principles. We intend to automate the Fidelity Index by developing rating scales to be applied automatically at critical decision-points for ePrescribing in individual patients using data from the EPMA system.

### Patient and public involvement

Two patient and public involvement (PPI) collaborators (AC, JB) have reviewed and commented on the plans and contributed to the study design, and have advised on the lay summary of the research.

### ePAMS+ intervention (figure 1)

Our intervention builds on the work of ARK (Antibiotic Reduction and Konservation), which produced an Antibiotic Review Kit that increased the proportion of antibiotic prescriptions reviewed within 72 hours from 91% to 99% and the proportion of antibiotic prescriptions stopped within 72 hours from 9% to 35%.^15^

**Figure 1.**
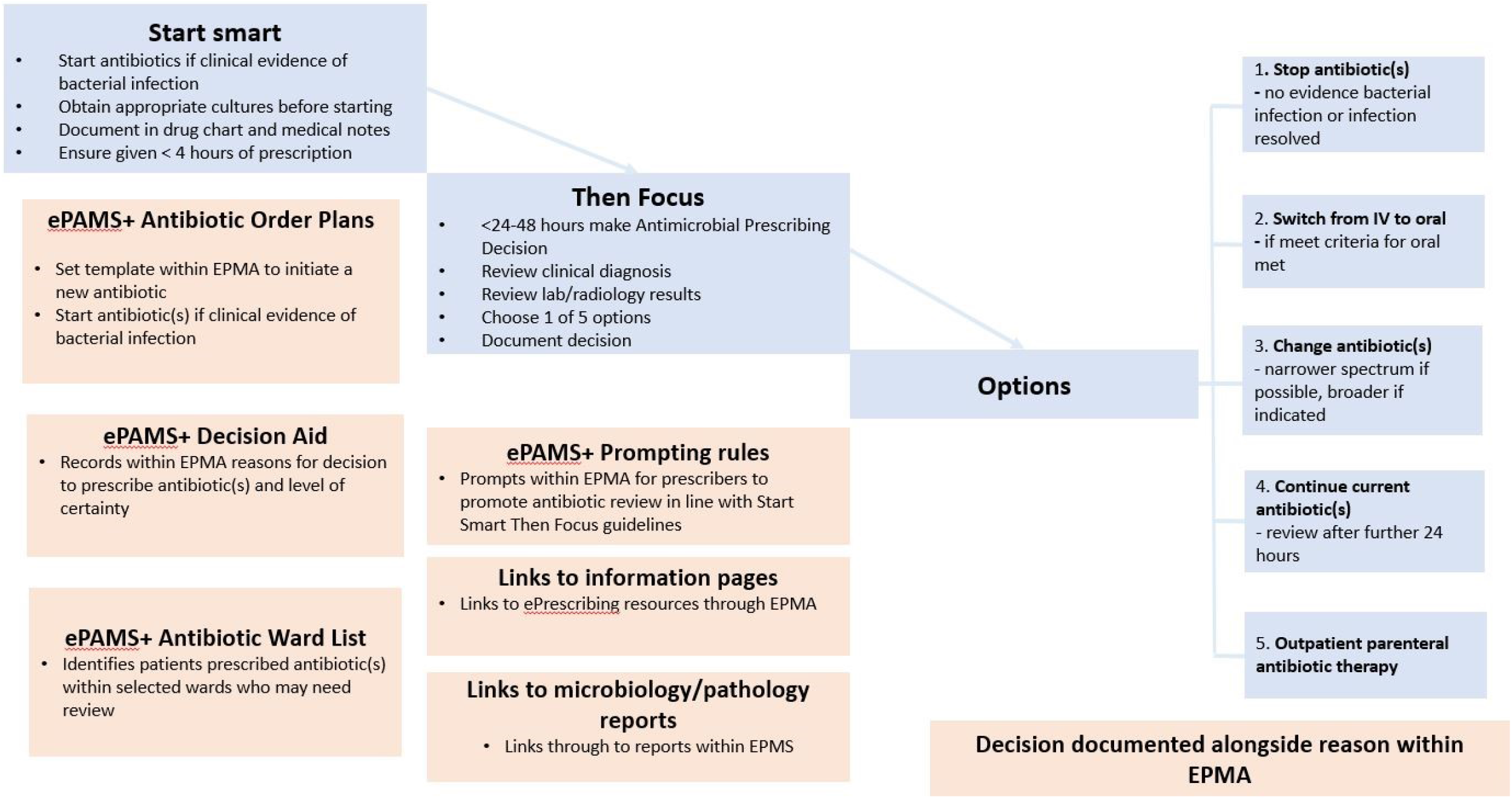
ePAMS+ intervention components alongside the national ‘Start Smart-Then Focus’ Guidelines.

ePAMS+ takes valuable lessons from the behavioural and organisational intervention work of ARK and extends it in three crucial ways:

1. Whereas ARK focused on stopping antibiotic prescribing at review, ePAMS+ aims to improve the decision-making process for all viable options, including starting and stopping treatment, optimisation of the dose regimen, switching the route of administration, changing the antibiotic, and continuing treatment for an appropriate duration;
2. ARK was implemented only in acute admission contexts whereas ePAMS+ applies to all hospital in- patient antibiotic prescribing; and
3. ARK was a behavioural and organisational intervention, whereas in addition to these aspects ePAMS+ also implements a CDS tool that exploits existing ePrescribing system functionality in order to automate, sustain and integrate effective support for appropriate antibiotic prescribing into all hospital prescribing pathways across multiple sites.

To inform the novel elements of ePAMS+, we have liaised extensively with policymakers, professional and patient representatives, vendors and international experts to conceptualise a prototype complex intervention which has the potential to support healthcare professionals and clinical teams at all key stages of antibiotic medicines management.^13^ We have identified the core requirements of ePAMS+ and how it can interface with Cerner Millennium, a commonly used commercial and integrated EPMA system in the UK. The principles of ePAMS+ have been designed to be adaptable for implementation in other EPMA systems.

Supplementary table 1 summarises the ePAMS+ intervention using the Template for Intervention Description and Replication (TIDieR) checklist.^16^ It aligns (figure 1) with best clinical practice and the national ‘Start Smart–Then Focus’ guidelines.^9^ These guidelines state that antibiotics should be started promptly for patients if there is a suggestion of bacterial infection; reviewed regularly within 48-72 hours of initial prescription to see if antibiotics are still needed; and stopped or switched as appropriate, once all test results to inform decision-making are received. The ePAMS+ intervention consists of the following tools embedded within the Cerner EPMA system:

- Antibiotic Order Plans (figure 2A&2B) to help prescribe antibiotics and schedule a series of review points (figure 2C) where changes in prescription may be required
- Decision Aid to help communicate the original prescriber’s level of certainty about the need for antibiotics in order to facilitate a later decision to cease prescription where appropriate (based on the ARK intervention classification^15^ of possible risk of infection, probable infection or finalised diagnosis of infection)
- Decision Aid (figure 2D) includes fields to record proposed site of infection (Body System) and working diagnosis (Indication)
- Information pages to help adopters benefit from using ePrescribing tools
- Antibiotic Ward Task List to identify patients on antibiotics that may need review
- Prompting rules for prescribers to promote antibiotic review
- Links to microbiology and/or pathology results within the review screen.

Within each participating hospital, an ePAMS+ Champion will form a local Implementation Team (see table 1 for details and approximate implementation timeline) to promote ePAMS+ through grand rounds, departmental/specialty team meetings, clinical governance meetings and training sessions for junior doctors/nurses/pharmacists. Prescribers, pharmacists and nurses working within study hospital wards will be encouraged to complete the ePAMS+ online eLearning training module.

**Table 1.** Implementation timeline.

| Timeline | Phase | Milestone |
| --- | --- | --- |
| Following site approval | Set-up activity | - Training of local ePAMS+ Champion<br>- Training of local ePAMS+ Implementation Team (comprising an AMS Lead; an antimicrobial pharmacist; a microbiologist and/or infection specialist; a medication safety officer; senior representatives from clinical areas impacted [including an acute/general consultant clinician who will act as an ePAMS+ Clinical Team lead]; a senior member of nursing staff; a specialty trainee doctor in year 3 or above; a core medical trainee; and a foundation doctor)<br>- Ensure Cerner EPMA system is ready to go live with ePAMS+ tools on the implementation/'go live' date<br>- Extract of pre-intervention dataset from Cerner EPMA system |
|  | Kick-off meeting preparation | - ePAMS+ Champion & Implementation Team: Finalise list of key members of clinical team<br>- ePAMS+ Champion & Implementation Team: Organise and publicise kick-off meetings |
| -2 Weeks | Kick-off and staff training | - ePAMS+ Implementation Team: Organise completion of ePAMS+ online tool training by clinical staff within selected areas<br>- ePAMS+ Champion & Implementation Team: Run kick-off meetings<br>- ePAMS+ Champion & Implementation Team: Organise regular, supportive discussion meetings with clinical team within selected areas |
| Week 1 | Implementation Go Live | - Activation/'go live' of ePAMS+ intervention within Cerner EPMA |
| Week 2+ | Implementation and data extraction | **Monitoring and Regular discussion of the implementation**<br>- Monthly extract of interim de-identified dataset from Cerner EPMA system<br>- Fidelity Index development- ePAMS+ Champion & Implementation Team: Undertake data collection on implementation of ePAMS+<br>-All Staff: Undertake regular, supportive discussion of the implementation of ePAMS+ (based on the latest monitoring data). |
| Week 16 | Assessment of long-term sustainability | - Extract and analysis of final dataset from Cerner EPMA system<br>- Fidelity Index completion<br>- Analysis of Feasibility Trial Results and Development of full-scale trial protocol |

**Figure 2.**
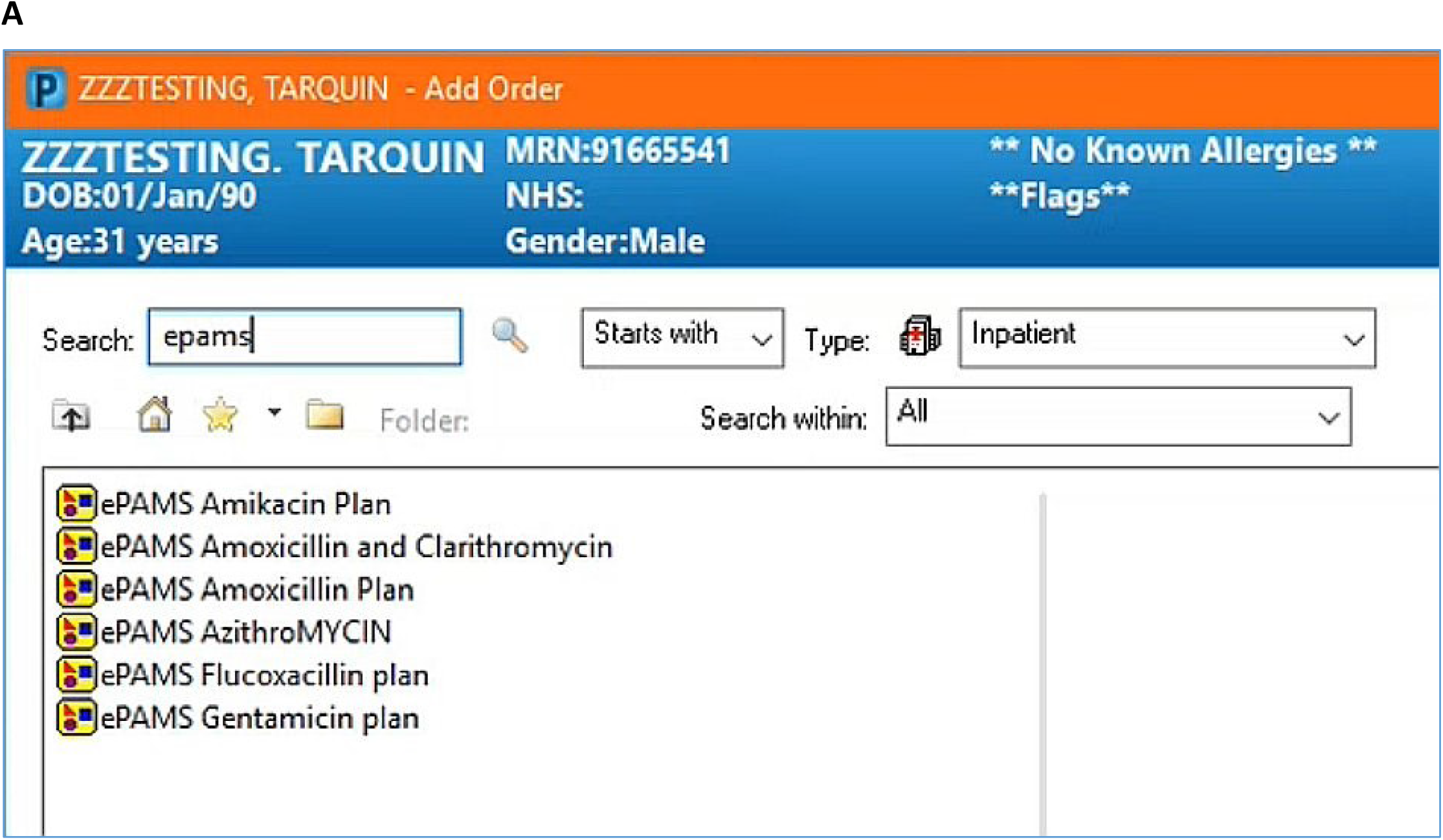
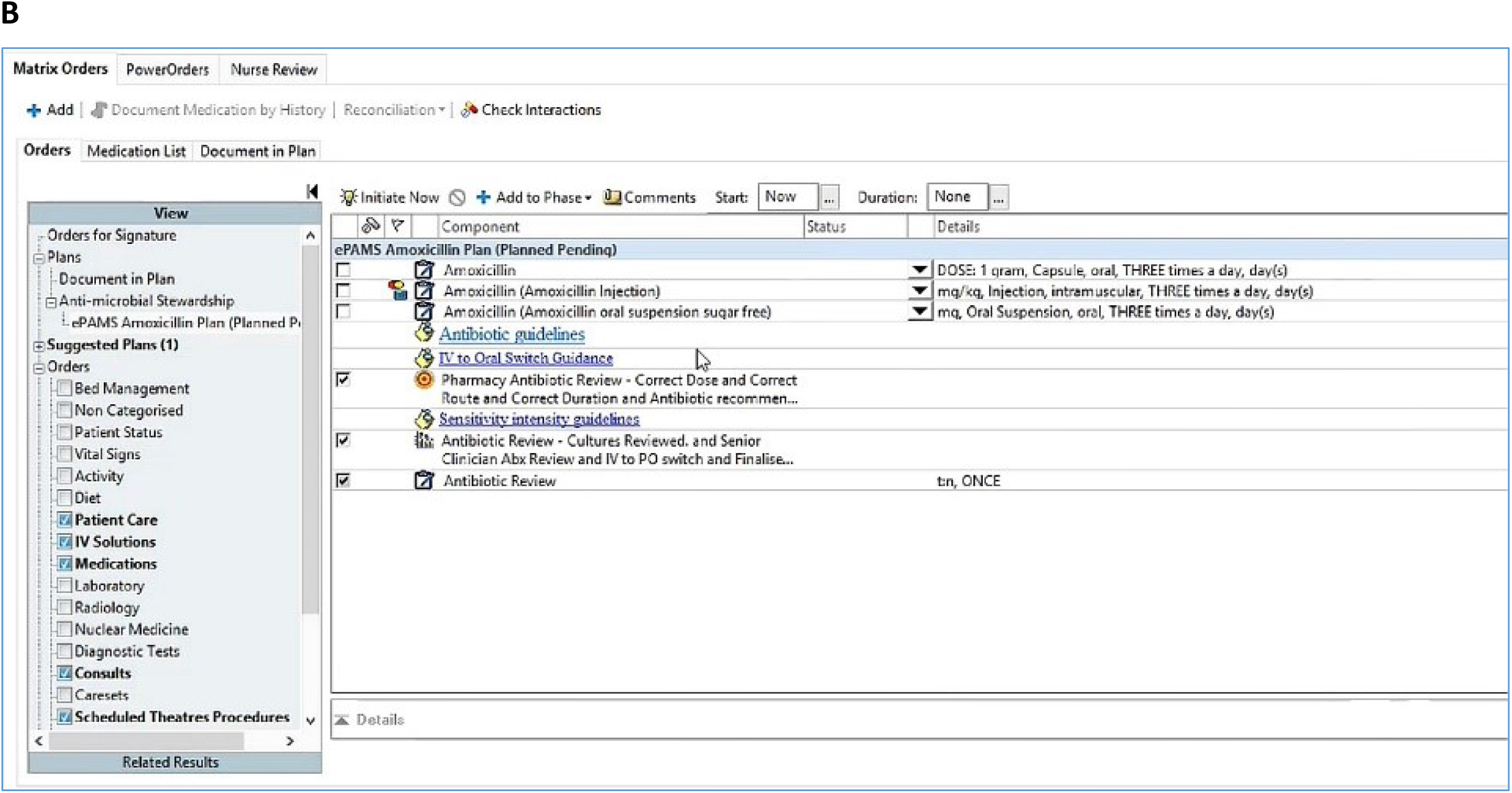
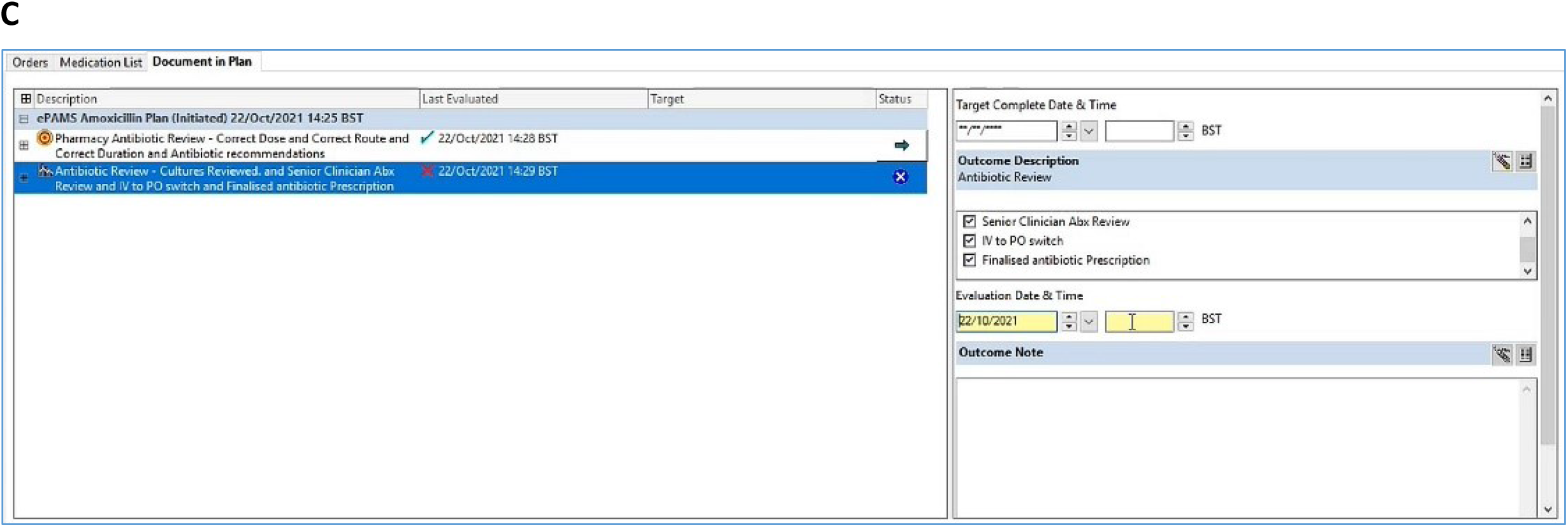
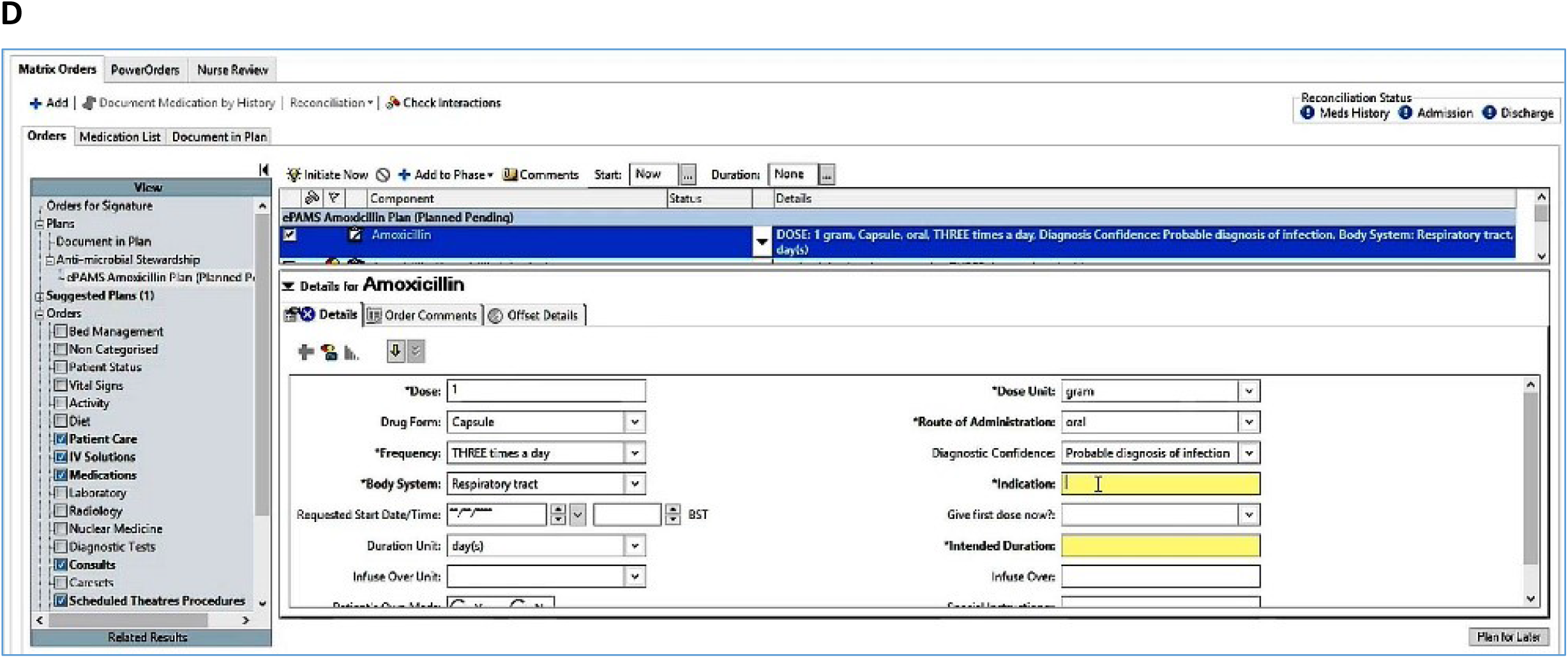
Screenshots illustrating technical components of the ePAMS+ intervention. A: ePAMS+ Order Plans B: ePAMS Amoxicillin Order Plan C: Antibiotic Review D: Decision Aid includes fields to record proposed site of infection (Body System) and working diagnosis (Indication)

### Setting

Two NHS Trusts which use the Cerner EPMA system have been selected as feasibility sites. Within each site, up to five wards will be purposively selected to ensure the ePAMS+ intervention feasibility is evaluated across a wide range of clinical settings.

### Qualitative component

Qualitative assessments will collect data in one multidisciplinary focus group at each site, approximately 10 hours of observation of clinical practice and approximately 10 interviews (five at each site). We will purposively sample a range of stakeholders. Participants will include both patients and staff including junior or senior doctors from a range of wards and specialties, nurses, pharmacists, IT and informatics staff, managers, other relevant healthcare professionals and system vendors. Staff participants may be involved in focus groups, interviews, and think-after interviews. Patients may participate in focus groups or interviews. Each focus group will include approximately 10-12 participants, incorporating up to three patients and up to nine staff participants. We anticipate that these numbers will lead to data saturation, giving us insights into potential intervention modifications to achieve maximum effectiveness and ensure acceptability to a range of stakeholders.

### Qualitative: patient participants

Patients will be eligible if they are able to provide informed consent, are aged ≥18 years, received antibiotic treatment in the last six months while in hospital, and are fluent in English. They will be excluded if they are temporarily unavailable (e.g., sleeping or receiving treatment) or if ward staff consider them too unwell to be interviewed. Potential participants will be identified by members of the direct care team according to the inclusion/exclusion criteria. An information poster for patients will also be displayed in wards.

### Qualitative: staff participants

Staff will be eligible if they are able to provide informed consent, are aged 18 years and older, have experience in dealing with prescribing or administration of antibiotics or ePrescribing systems and are either junior or senior doctors, nurses, pharmacists, IT staff, managers, other relevant healthcare professionals, or vendors.

Staff will be approached via two pathways, either on recommendation of a senior clinician on their ward or through recruitment leaflets displayed in wards. Individual participants at each site will either be approached in person (where feasible) or by telephone or email to enquire whether they are interested in participating in the study and if so whether they would prefer a face-to-face or telephone interview.

All potential patient and staff participants will receive written information on the project from the research team, outlining what participation will involve. They will be given at least 24 hours to consider their decision to participate and can withdraw at any point. Upon receiving the completed consent form, a researcher will contact participants in order to arrange a suitable time for an interview or focus group.

### Qualitative: data collection

#### i. Focus groups

Focus groups will take place remotely and will last no more than 60 minutes. Each focus group will cover experiences and opinions of ePAMS+ from a variety of perspectives, and explore its potential wider usability. Focus groups will be audio-recorded (if all participants agree) and transferred on encrypted equipment. If, however, audio-recording is not consented to by all participants, researchers will take detailed notes from the focus group session.

#### ii. Interviews

Interviews will be conducted online via Microsoft Teams or Zoom depending on local requirements and will be recorded using the online platform. All audio-recordings will be transcribed by an external transcription company contracted to the University of Edinburgh.

Interviews will be one-to-one and conducted remotely. Patients will be interviewed about their experiences of being prescribed antibiotics in hospital, and their thoughts around reducing inappropriate prescribing of antibiotics and the potential impact on them. Staff will be asked about the way they work, how they use IT systems, what they think about AMS and how it can be promoted. Interviews will take up to an hour each, but could be significantly shorter depending on participant preference. Participants may choose to take part in up to three interviews over the course of the project.

#### iii. Think-after Interviews

Staff who undergo think-after interviews will be asked to say aloud everything they were thinking about on each page while they were using the ePAMS+ tools. Think-after interviews will be one-to-one and conducted remotely and will take up to an hour each (but could be significantly shorter depending on participant preference). Participants may choose to take part in up to three think-after interviews.

#### iv. Observations

Staff who participate in observations will be shadowed by a researcher during their normal working day. The length of observations could range from 30 minutes to up to four hours, depending on participant preferences. During the observation, the researcher will take notes about their impressions of how the participant uses ePAMS+. Observations will be non-participant in nature.

### Qualitative: analysis

Qualitative data collection and analysis will be iterative, allowing emerging themes to be explored further and disconfirming evidence to be sought. Thematic analysis will allow us to access a diverse range of interviewees/perspectives, facilities and contexts. Detailed within-case analysis will be followed by analysis across cases to identify over-arching themes, similarities and differences between cases, and potential implications. Results of the analysis will inform development and implementation of the intervention.

Thematic analysis of focus group, interview and observation data will investigate how the intervention was received and how it may need to be adapted for other contexts and to identify likely mechanisms of action to be examined in the process evaluation in the future full-scale stepped-wedge trial evaluating the ePAMS+ intervention. Issues regarding effective design (usability; fit with existing workflows) and implementation (training; user acceptance) of the ePAMS+ intervention will be explored.

### Quantitative component

We aim to study at least 100 admissions per ward to enable precise estimation of between-patient variability, by ward and overall, in antibiotic use and to explore feasibility of data extraction across a wide range of clinical presentations. Furthermore, inclusion of a diverse range of wards and cases will support the development of the Fidelity Index quantifying the extent to which practice has adhered to ePAMS+ core principles.

### Quantitative: patient participants

Individuals eligible for inclusion in the study will be aged ≥16 years, will have been admitted to hospital as a medical inpatient and will have an antibiotic order plan initiated or an existing antibiotic prescription flagged within the EPMA.

As ePAMS+ is a service-level intervention, all eligible admissions to participating wards in study sites will be included in quantitative analyses. Although patient informed consent is not required or sought as part of this study, the implementation pack contains a patient information leaflet to help clinical staff explain the process of antibiotic use and review to patients. There is no mechanism to allow patients within participating wards from opting out of the collection and use of routine de-identified administrative data.

### Quantitative: data collection

Under the ePAMS+ intervention, prescription of antibiotics automatically flags within the EPMA system to trigger decision aids and task lists for appropriate antibiotic management. Information on how order plans, prescribing interventions and review processes are managed within the Cerner EPMA system using the ePAMS+ tool is available in the supplementary information on the intervention (supplementary table 1).

Data on EPMA interactions will be automatically logged within the data audit system of the Cerner EPMA system already in use at trial sites. A standardised data query to the system will be run by the local NHS Trust information services team. These queries will be run regularly within the data audit system (prior to activation of the ePAMS+ intervention and at intervals after implementation). We will extract two types of data: outcomes for quantitative analysis purposes, such as data contributing to calculation of total antibiotic use; and process measures (table 2) to help understand how the ePAMS+ system is being used.

**Table 2.**
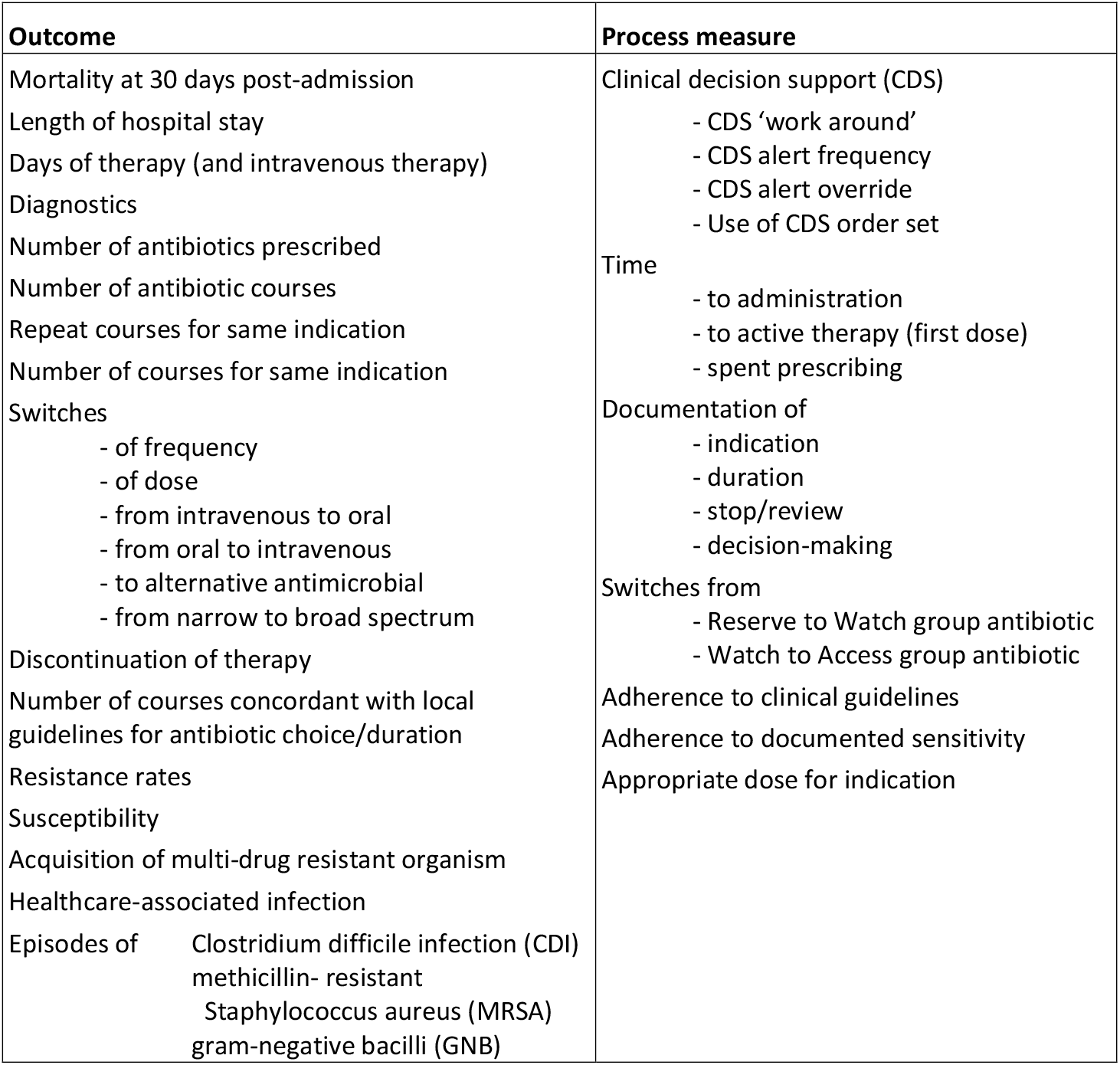
Feasibility trial quantitative secondary outcomes and process measures.

Personal data will be processed as follows. Prior to data extraction from the Cerner EPMA system, a unique non-identifiable alias will be created for each record. The data extracted will not include any direct identifiers, but will include participant age at time of extract, diagnosis and ward of treatment, in addition to details of antibiotic prescriptions received. Data extracted from participating NHS Trusts will be transferred via secure file transfer protocol (Serv-U FTP) to the National Safe Haven maintained by Public Health Scotland. Data controller/data controller information sharing agreements will be established between each site and University of Edinburgh (Sponsor).

All data will be held in a project-specific area in the National Safe Haven maintained by Public Health Scotland with access limited to named project researchers via a unique username and multi-factor authentication. All use will be subject to a user agreement covering responsibilities, access requirements, data security and processes for release of analytical output. The National Safe Haven will review all outputs to ensure these would not disclose the identity of any participant.

### Quantitative: training and learning data

Site staff ePAMS+ training information will be captured on the Learning Management System to assess completion of training (i.e. professional discipline, date/time of module completion, time spent on learning and pre and post-test scores).

### Quantitative: outcomes

We will assess feasibility of standardised queries to capture data from local Cerner EPMA system configurations and completeness of the data extracted. Further important feasibility assessments will be the ability to derive total antibiotic consumption, measured as the number of defined daily doses (DDD) per admission, and to obtain from local hospital systems mortality at 30 days post-admission; these will be co-primary outcomes for the future full-scale trial. Table 2 outlines the other outcomes and process measures to be gathered.

A data will be extracted at intervals and the process of extraction is a key feasibility objective, there will not be scope to monitor the occurrence of adverse events in real time. All patients within participating sites will be managed according to best clinical practice and in line with local and national clinical guidelines.

### Quantitative: analysis

Descriptive summary statistics will be reported on antibiotic consumption, overall and by site. Separate summaries will also be provided for intravenous, oral, broad spectrum and narrow spectrum antibiotics.

Between-patient variability in total antibiotic consumption, measured as the number of DDD per admission, will be quantified using a normal linear model, including site and ward as factors, to estimate the components of variance. Log-transformation will be performed if necessary to satisfy the model assumptions. Factors for seasonal effects and implementation of the ePAMS+ intervention will also be considered.

Other quantitative outcomes (table 2) will be assessed according to two criteria. First, we will determine whether it is possible to derive each outcome using the information available in the EPMA data extract. Secondly, we will summarise the measures descriptively, overall and by site and by ward, with a particular focus on the rate of missing data.

### Quantitative: Fidelity Index

A Fidelity Index will be developed to capture the extent to which prescribers apply ePAMS+ ‘core principles’ (e.g. using the decision-aid for review and revise or using the patient leaflet for shared-decision making), in their practice. Assessing fidelity helps increase confidence that changes in the dependent variable are attributable to the independent variable and that behavioural interventions are implemented as described in the protocol.^17 18^

Through Cerner EPMA system data extracts we will:

#### i. Explore the critical decision-making points for prescribers

We will map behavioural elements of ePAMS+ to the data, to identify which items must be tagged for automation. One such element would be the diagnostic confidence decision aid, in which the prescriber rates their certainty about the presence of infection: none; possible risk from infection; probable diagnosis of infection; prophylaxis. The corresponding data item would be whether diagnostic confidence had been recorded in the EMPA at initial prescription. These behavioural elements will be the ‘critical decision-making points’ for prescribers that reflect the application of the ePAMS+ intervention core principles in their practice. This is critical for outcomes evaluation in the future full-scale trial, as these items would serve as ‘intermediate outcomes’ to help explain the relationship between the outcome and the intervention.

#### ii. Understand the data structure for automating the fidelity coding

We will identify the critical decision-making points for prescribing within Cerner EPMA (relevant to ePAMS+), develop codes for automatic categorisation of their level of implementation and consider key locations within an EPMA system where these can be embedded.

#### iii. Develop individual and composite scales for capturing practice

This part of the Fidelity Index measures the ‘actual’ implementation of ePAMS+ intervention as opposed to the ‘intended’. This will involve quantifying each ‘critical decision-making point’ (for example, whether antibiotic review was conducted within 48-72 hours of initial prescription) into a 3- point rating scale that reliably discriminates between ‘fully’, ‘partially’ and ‘not implemented’. Although successfully used previously,^19-21^ these categories might not apply to ePAMS+. The feasibility work will help confirm these categories or explore alternatives such as codifying into ‘present’, ‘absent but should be present’ and ‘not applicable’.^22^ The scores from the rating scales will combine in a cumulative score for intervention fidelity (per case, per prescriber) for linking with outcome measures.

After developing the Fidelity Index, the specifications for its automation within Cerner EPMA systems and the methods of deriving summary measures of antimicrobial use, these will be pre-tested in the feasibility trial.

### Progression to larger-scale trial

At the end of the feasibility study the investigators will meet to review and integrate the quantitative and qualitative findings. They will decide whether the feasibility results merit progression to the planned larger-scale effectiveness trial. If a decision is made to progress the meeting will also generate a list of any proposed modifications to the ePAMS+ intervention and its implementation to be actioned prior to the larger trial commencing.

## Supporting information

SPIRIT checklist

## Data Availability

All data produced in the present study are available upon reasonable request to the authors, subject to ethics and information governance approval

## Ethics and dissemination

The qualitative research and Fidelity Index have been approved by the Health and Research Authority and the North of Scotland Research Ethics Service (ref:19/NS/0174). The feasibility trial and quantitative analysis have been approved by the London South East Research Ethics Committee (ref:22/LO/0204). An independent trial steering committee, composed of members of the independent programme steering committee for grant RP-PG-0617-20009, will oversee the trial conduct.

Results will be shared with study sites and with participants in the qualitative research. Findings will be published in peer-reviewed journals and presented at academic conferences. Published results will not contain any personal data and will be in a form where individuals are not identified and re-identification is unlikely to take place.

## Contributorship statement

CJW, KC and RW conceived this paper and CJW led the drafting of the manuscript. AH and NW led the development of the intervention under the oversight of the ePAMS+ Programme Management Group. ASh is the PI of the programme and oversees all aspects of the research. All authors reviewed and commented on drafts of the manuscript.

## Acknowledgements

We gratefully acknowledge the input of the wider team of ePAMS+ researchers, staff and co-investigators, and the Independent Steering Committee for its oversight of this programme.

