## Supplemental table 1 for "A complex ePrescribing-based Anti-Microbial Stewardship (ePAMS+) intervention for hospitals combining technological and behavioural components: protocol for a feasibility trial"

**Table 1** **Description of the ePAMS+ intervention according to the TIDieR checklist**

| Item Number | Item Description |
|---|---|
| | **BRIEF NAME OF INTERVENTION** |
| **1.** | ePrescribing-based Anti-Microbial Stewardship Intervention for Hospitals (ePAMS+) |
| | **WHY** |
| **2.** | Reducing the risk of antimicrobial resistance through appropriate antibiotic use is a major national and global challenge. The UK Department of Health recommends a 'review and revise' approach to antibiotic prescribing in hospitals ('Start Smart – Then Focus'). Within this approach antibiotics should be started promptly for patients who have a possible bacterial infection, reviewed regularly within the first 24-72 hours of initial prescription to see if antibiotics are still needed (based on cultures and sensitivities, test results and the patient's clinical presentation) and then stopped or switched if no longer needed.<br><br>Despite national guidance, antibiotic prescribing rates are still increasing in UK hospitals. The reasons are multi-factorial: including limited real-time access to information that may influence decisions to initiate antibiotics, concerns about missing potentially serious infection (e.g. sepsis), and lack of continuity of care, which limits the opportunity for an informed review of progress and results in a reluctance to delay initiation of an/or change/discontinue an antibiotic that has already been commenced. The substantial time pressures that clinicians face compound these challenges.<br><br>The UK National Institute for Health Research (NIHR) funded programme developed a complex ePrescribing-based Anti-Microbial Stewardship (ePAMS+) intervention that aligns with the UK national strategy and aims to safely reduce inappropriate antibiotic use in adult medical in-patient populations. |
| | **WHAT** |

**3.** ePAMS+ is an intervention developed to align with the national 'Start Smart – Then Focus' guidelines. It consists of a series of ePrescribing tools designed to embed within the Cerner ePrescribing and Medication Administration (EPMA) system:

- Antibiotic Order Plans to help prescribe antibiotics and set up review and revise processes
- Decision Aid to help communicate the original prescriber's level of certainty about the need for antibiotics (based on ARK intervention)
- Information pages within the EPMA to help team get most from ePrescribing tools when used
- Antibiotic Ward Task List to identify patients on antibiotics that may need review
- Decision aid will include fields to record proposed site of infection (Body System) and working diagnosis (indication)
- Prompting rules for prescribers to promote antibiotic review
- Links to microbiology results +/- pathology results within the review screen

Materials

- Patient Information Leaflet
- Powerpoint presentation: Description of each slide:

Slide 1: What is ePrescribing-based Anti-Microbial Stewardship Plus (ePAMS+)?
Slide 2: Start Smart - Then Focus guidelines
Slide 3: What are the risks of staying on antibiotics longer than needed?
Slide 4: How will ePAMS+ help?
Slide 5: What tools does ePAMS+ provide?
Slide 6: Antibiotic Order Plans
Slide 7: How do I find the ePAMS+ Order Plans
Slide 8: What do Order Plans look like?
Slide 9: Using the Order Plan
Slide 10: Using the Order Plan – review and optimisation
Slide 11: Using the Order Plan – cessation/switching
Slide 12: ARK Decision Aid
Slide 13: The ARK Decision Aid categories
Slide 14: How can I use the ARK Decision Aid in ePAMS+
Slide 15: Patient Leaflet
Slide 16: Antibiotics ward task lists
Slide 17: Information pages in ePrescribing
Slide 18: Data collection and regular, supportive team meetings

Slide 19: What next?

Implementation guidance

Website: www.ed.ac.uk/usher/research/projects/epams

Training video demonstration, with transcript, of ePAMS Prescribing Tool

Describe any physical or informational materials used in the intervention, including those provided to participants or used in intervention delivery or in training of intervention providers. Provide information on where the materials can be accessed (e.g. online appendix, URL).

**4.**

- Implementation guidance in the form of a written manual and online website

When a patient is prescribed antibiotics (either a single drug or combination) a prompt will appear to place the patient on review and a decision tool will record the reason for the choice of antibiotic treatment. If antibiotic treatment is chosen on a 'just in case' basis then prescribers [or other clinicians involved in patient care?] are asked to review antibiotic treatment in the following ways, in line with the national guidelines:

- Patients on review have antibiotics stopped if clinicians decide that their illness is not caused by bacteria
- Patients on review have antibiotic course duration and/or type changed by clinicians depending on results from test results
- Patients on review have antibiotics continued because clinicians decide they are right for their illness

**WHO PROVIDED**

5.

- Within each participating hospital, an ePAMS+ ‘Champion’ will be identified to promote the intervention.
- The local ePAMS+ Champion will form an Implementation Team comprising antimicrobial stewardship (AMS) Lead, an antimicrobial pharmacist, a microbiologist and/or infection specialist, a medication safety officer, senior representatives from clinical areas impacted (which should include an acute/general consultant clinician who will act as an ePAMS+ Clinical Team lead), a senior member of nursing staff, a specialist trainee (ST3+), a core medical trainee (CMT) and a foundation doctor.
- Key requirements: the ePAMS+ intervention team should be familiar with the difference between a probable diagnosis of infection versus a possible risk of infection, and the evidence of lack of harm from shorter antibiotic courses; discuss how to integrate the ePAMS+ Decision Model within local practice; importance of first reviewer communicating their degree of confidence to the reviewer; establish whether review data available on review and revise – or an audit to see baseline levels of review and revise; identify and discuss potential barriers and solutions; plan launch and establishment of ePAMS+
- Implementation team tasked with promoting ePAMS+ through the channels available to them: grand round, departmental/specialty team meetings, clinical governance meetings, teaching sessions for junior doctors/nurses/pharmacists i.e. generic skills, mandatory training sessions
- Completion of ePAMS+ online training by all prescribers, pharmacists and nurses working within selected hospital areas

**HOW**

**6.** Initial training of the ePAMS+ Implementation Team would be via remote or face-to-face meeting with online access set up to a training tool/slide deck. Training would be via the online training with face-to-face meetings to reinforce/promote use wherever possible.

**WHERE**

**7.** Describe the type(s) of location(s) where the intervention occurred, including any necessary infrastructure or relevant features.

- UK hospital adult in-patient wards using the Cerner EPMA system, excluding surgical wards.
- UK hospital pharmacies.

---

**WHEN and HOW MUCH**

**8.** Describe the number of times the intervention was delivered and over what period of time including the number of sessions, their schedule, and their duration, intensity or dose.

Within selected wards, the ePAMS+ intervention would be delivered every time an antibiotic prescription is made (either for a single drug or combination) and can be used to manage the full duration of each prescription.

**TAILORING**

**9.** N/A

**MODIFICATIONS**

**10.‡** N/A

**HOW WELL**

**11.** Planned: If intervention adherence or fidelity was assessed, describe how and by whom, and if any strategies were used to maintain or improve fidelity, describe them.

The feasibility trial will incorporate development of a Fidelity Index for use in the future full-scale evaluation of ePAMS+.

**12.‡** Actual: If intervention adherence or fidelity was assessed, describe the extent to which the intervention was delivered as planned.

Usual antibiotic prescribing practice in study sites will constitute the comparator group in the trial. There will be no restriction on concomitant care and interventions during the course of the trial. No formal application of the Fidelity Index will be used in the feasibility study, as it will be developed as part of this trial. Qualitative studies of implementation will give an indication of the extent to which the intervention was delivered as planned.

---

N/A: item not applicable for the intervention being described.

‡ If completing the TIDieR checklist for a protocol, these items are not relevant to the protocol and cannot be described until the study is complete.
